# Prevalence and Impact of Myocardial Injury in Patients Hospitalized with COVID-19 Infection

**DOI:** 10.1101/2020.04.20.20072702

**Authors:** Anuradha Lala, Kipp W Johnson, James Januzzi, Adam J Russak, Ishan Paranjpe, Shan Zhao, Sulaiman Somani, Akhil Vaid, Fayzan Chaudhry, Jessica K De Freitas, Felix Richter, Zahi A Fayad, Sean P. Pinney, Matthew Levin, Alexander Charney, Emilia Bagiella, Jagat Narula, Benjamin S Glicksberg, Girish Nadkarni, Donna M. Mancini, Valentin Fuster, on behalf of the Mount Sinai Covid Informatics Center

**Author notes:** Correspondence: Anuradha Lala, 1 Gustave Levy Place, Box 1030, New York, NY 10029, (P), (F). These authors contributed equally (Anuradha Lala, Kipp W Johnson). These authors contributed equally and jointly supervised this work (Donna M. Mancini, Valentin Fuster).

## Abstract

**Background:** The degree of myocardial injury, reflected by troponin elevation, and associated outcomes among hospitalized patients with Coronavirus Disease (COVID-19) in the US are unknown.

**Objectives:** To describe the degree of myocardial injury and associated outcomes in a large hospitalized cohort with laboratory-confirmed COVID-19.

**Methods:** Patients with COVID-19 admitted to one of five Mount Sinai Health System hospitals in New York City between February 27th and April 12th, 2020 with troponin-I (normal value <0.03ng/mL) measured within 24 hours of admission were included (n=2,736). Demographics, medical history, admission labs, and outcomes were captured from the hospitals’ EHR.

**Results:** The median age was 66.4 years, with 59.6% men. Cardiovascular disease (CVD) including coronary artery disease, atrial fibrillation, and heart failure, was more prevalent in patients with higher troponin concentrations, as were hypertension and diabetes. A total of 506 (18.5%) patients died during hospitalization. Even small amounts of myocardial injury (e.g. troponin I 0.03-0.09ng/mL, n=455, 16.6%) were associated with death (adjusted HR: 1.77, 95% CI 1.39-2.26; P<0.001) while greater amounts (e.g. troponin I>0.09 ng/dL, n=530, 19.4%) were associated with more pronounced risk (adjusted HR 3.23, 95% CI 2.59-4.02).

**Conclusions:** Myocardial injury is prevalent among patients hospitalized with COVID-19, and is associated with higher risk of mortality. Patients with CVD are more likely to have myocardial injury than patients without CVD. Troponin elevation likely reflects non-ischemic or secondary myocardial injury.

**Unstructured Abstract:** Myocardial injury reflected as elevated troponin in Coronavirus Disease (COVID-19) is not well characterized among patients in the United States. We describe the prevalence and impact of myocardial injury among hospitalized patients with confirmed COVID-19 and troponin-I measurements within 24 hours of admission (N=2,736). Elevated troponin concentrations (normal <0.03ng/mL) were commonly observed in patients hospitalized with COVID-19, most often present at low levels, and associated with increased risk of death. Patients with cardiovascular disease (CVD) or risk factors for CVD were more likely to have myocardial injury. Troponin elevation likely reflects non-ischemic or secondary myocardial injury.

## INTRODUCTION

Coronavirus Disease (COVID-19), caused by severe acute respiratory syndrome coronavirus-2 (SARS-CoV-2), is now one of the deadliest pandemics in modern history. The mode of infection of COVID-19 is thought to be direct entry of the SARS-CoV-2 virus into cells via the human angiotensin-converting enzyme 2 (ACE2) receptor, which is expressed predominantly in the lungs but also throughout the cardiovascular system (1). Thus, while the most virulent manifestation of COVID-19 is acute respiratory distress syndrome (ARDS), select reports from Europe and China have also demonstrated cardiac injury reflected through elevated troponin concentrations among infected patients (2-5). In these limited case series, troponin elevation was more common in patients with preexisting cardiovascular disease and, when present, was associated with higher rates of adverse outcomes in patients hospitalized with COVID-19 (6). However, the observational nature and small sample sizes limit the generalizability of these findings. Additionally, there are no large studies from the United States (US), the current epicenter of the pandemic.

As such, major gaps remain in our current understanding of the underlying mechanisms by which SARS-CoV-2 affects the cardiovascular system and how such involvement may alter clinical outcomes: First, the range of troponin elevation across different subpopulations based on history of cardiovascular disease (CVD) compared to those without history of CVD is unknown among patients in the US. Second, whether these troponin elevations represent primary myocardial infarction, supply-demand inequity, or non-ischemic myocardial injury remains unclear. Finally, the impact of myocardial injury in the context of COVID-19 infection on outcomes is not well studied. We sought to explore these aims amongst a large cohort of patients hospitalized with COVID-19 in New York City.

## METHODS

### Study Population

Patients in this study were drawn from five New York City hospitals (Mount Sinai Hospital, in East Harlem; Mount Sinai West, in Midtown Manhattan; Mount Sinai St. Lukes, in Harlem; Mount Sinai Queens, in Astoria; and Mount Sinai Brooklyn, in the Midwood neighborhood of Brooklyn). These hospitals comprise the Mount Sinai Health System (MSHS). We included all patients admitted to a MSHS hospital with a laboratory confirmed SARS-CoV-2 infection who were at least 18 years old and had a troponin measurement within the first 24 hours of admission between February 27th and April 12th, 2020. The Mount Sinai Institutional Review Board approved this research under a regulatory protocol allowing for analysis of patient-level COVID-19 data.

### Data Collection

Data was collected from electronic health records (EHR) from the five hospitals. Variables collected included demographics, laboratory measurements, disease diagnoses, comorbidities, procedures, and outcomes (death or hospital discharge). Comorbidities were extracted using International Classification of Disease (ICD) 9/10 billing codes for atrial fibrillation (AF), asthma, coronary artery disease (CAD), cancer, chronic kidney disease (CKD), chronic obstructive pulmonary disease (COPD), diabetes (DM), heart failure (HF), and hypertension (HTN). Troponin I concentrations were assessed via the Abbott Architect method (Abbott, Abbott Park, Illinois) wherein the 99th percentile for a normal population is 0.028 ng/mL. The reference level for normal in MSHS is less than 0.03 ng/mL.

### Statistical Analysis

Descriptive analyses were performed by troponin levels stratified into normal (0.00-0.03 ng/mL), mildly elevated (between one and three times the upper limit of normal, or >0.03-0.09 ng/mL), and elevated (more than three times the upper limit of normal, or >0.09 ng/mL). Categorical variables were reported as total count and percentage of patients. Continuous non-troponin laboratory values were reported as median and interquartile range. We used troponin measurements within 24 hours of admission. If multiple troponin measurements were available within 24 hours, the patient’s first measurement was used.

To assess the effects of troponin levels on outcomes, we conducted a survival analysis with the dependent variable of time to mortality, setting time zero at time of hospital admission. Patients were considered to be right-censored if they were (1) discharged from the hospital alive or (2) remained in the hospital at the time of data freeze (April 12th). We fit Cox proportional hazards regression models with mortality as the dependent variable, adjusting for age, sex, race, ethnicity, history of CAD, history of AF, history of HF, history of HTN, history of CKD, history of DM, and in-hospital intubation. Age was modeled as age at time of admission, while gender and history of CAD, AF, HTN, CKD, DM, and in-hospital intubation were modeled with binary variables. Self-declared race was included in the model stratified in indicator variables corresponding to Caucasian, African American, Asian, Pacific Islander, Other, or Unknown with Caucasian as reference. Self-declared ethnicity was included in models as Hispanic/Latino, Non-Hispanic/Latino, and Unknown with Hispanic/Latino as the reference level. We tested for proportional hazards assumption by plotting Martingale residuals from the Cox proportional hazards model vs. linearized predictions. 95% confidence intervals from a LOESS best-fit line fit to the Martingale residuals included 0 for all values of the linear predictions, indicating visually there was not significant violation of the proportional hazards assumption. We then plotted Kaplan-Meier curves for survival stratified by troponin group. All analyses were conducted in R version 3.6.1, and visualizations were produced using the survminer R package (7,8).

For a sensitivity analysis, we conducted a secondary survival analysis where discharge from the hospital was considered to be a competing risk since mortality status could not be assessed after hospital discharge. We used the cmprsk R package for this analysis (9). Hazard ratios for the troponin variables from this analysis were not meaningfully different from our standard survival analysis.

## RESULTS

### Patient Characteristics and Troponin Levels

During the study period, 3,047 COVID-19 positive patients were hospitalized at one of five MSHS New York City hospitals. Of these, 2,736 (89.8%) had at least one troponin-I measurement within 24 hours of admission. The median age was 66.4 years, 40.7% of patients were over age 70, and 59.6% were male. One-quarter of all patients self-identified as African American and 27.6% self-identified as Hispanic or Latino. A history of CVD, including CAD, AF, or HF, was present in 35% of patients. Risk factors for CVD of DM or HTN, were present in 65% of the cohort (**Table 1**).

**Table 1.**
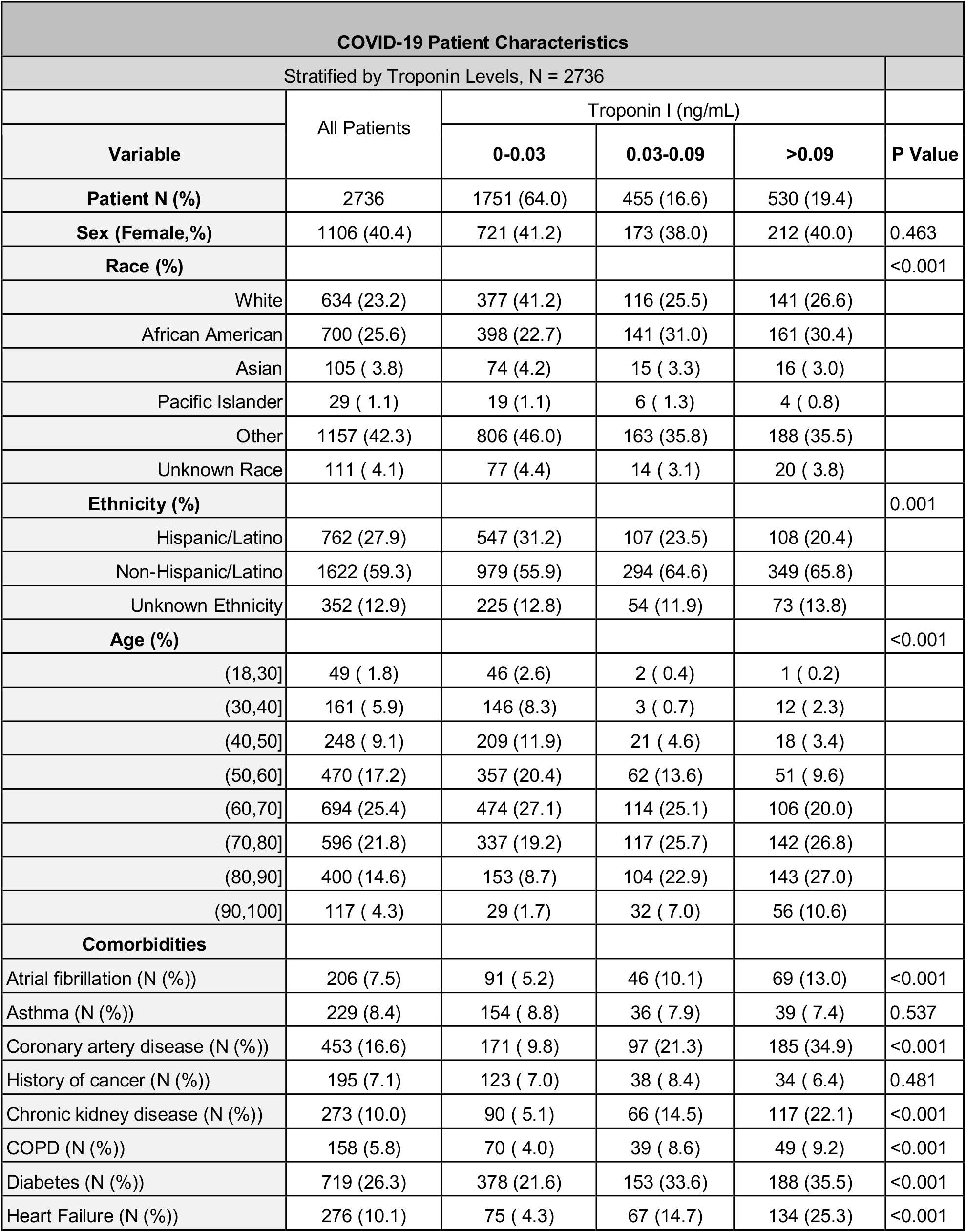
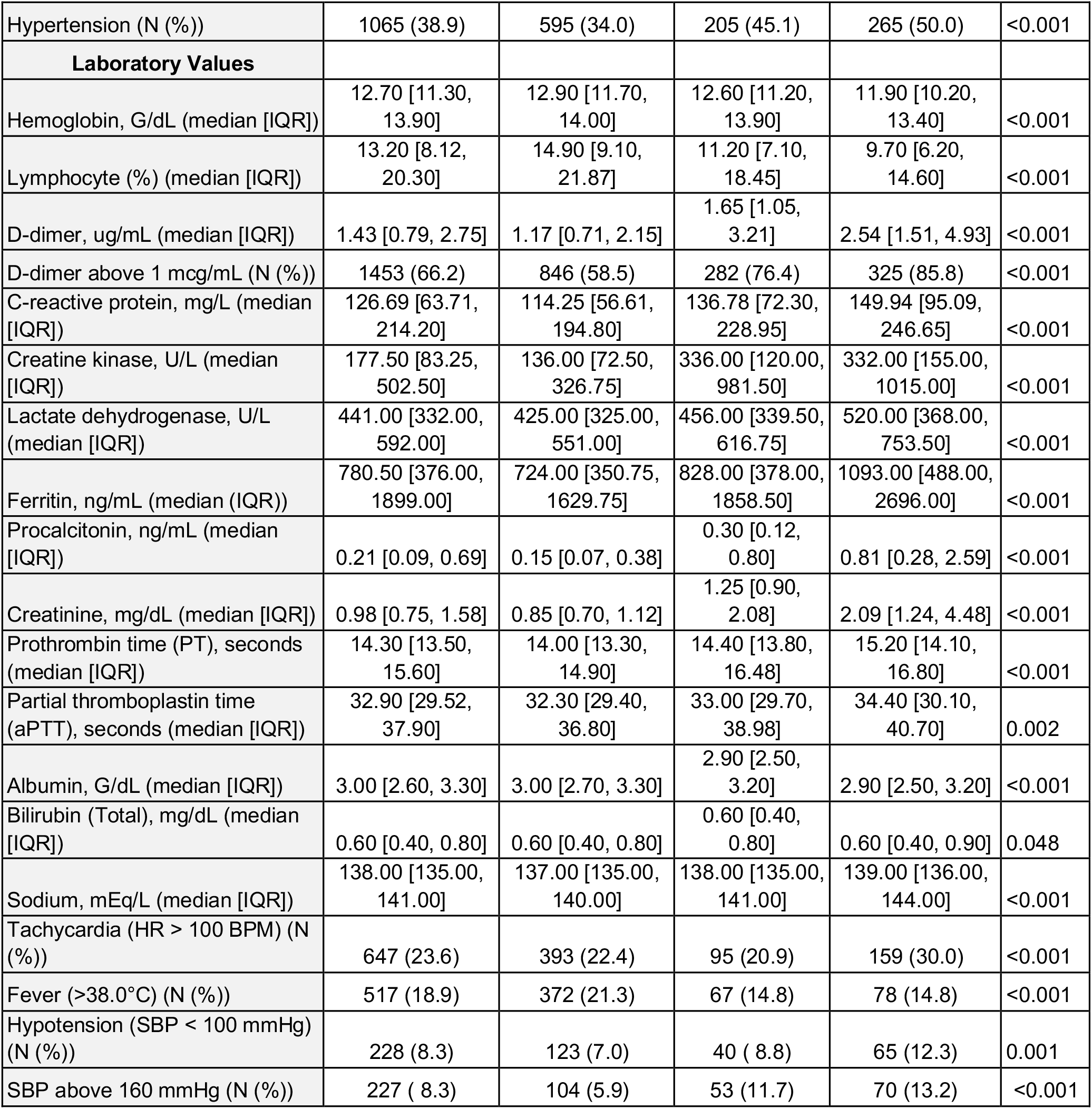
Baseline characteristics of admitted patients, stratified by troponin concentration.

| COVID-19 Patient Characteristics |  |  |  |  |  |
| --- | --- | --- | --- | --- | --- |
| Stratified by Troponin Levels, N = 2736 |  |  |  |  |  |
|  | All Patients | Troponin I (ng/mL) |  |  |  |
| Variable |  | 0-0.03 | 0.03-0.09 | >0.09 | P Value |
| Patient N (%) | 2736 | 1751 (64.0) | 455 (16.6) | 530 (19.4) |  |
| Sex (Female,%) | 1106 (40.4) | 721 (41.2) | 173 (38.0) | 212 (40.0) | 0.463 |
| Race (%) |  |  |  |  | <0.001 |
| White | 634 (23.2) | 377 (41.2) | 116 (25.5) | 141 (26.6) |  |
| African American | 700 (25.6) | 398 (22.7) | 141 (31.0) | 161 (30.4) |  |
| Asian | 105 ( 3.8) | 74 (4.2) | 15 ( 3.3) | 16 ( 3.0) |  |
| Pacific Islander | 29 ( 1.1) | 19 (1.1) | 6 ( 1.3) | 4 ( 0.8) |  |
| Other | 1157 (42.3) | 806 (46.0) | 163 (35.8) | 188 (35.5) |  |
| Unknown Race | 111 ( 4.1) | 77 (4.4) | 14 ( 3.1) | 20 ( 3.8) |  |
| Ethnicity (%) |  |  |  |  | 0.001 |
| Hispanic/Latino | 762 (27.9) | 547 (31.2) | 107 (23.5) | 108 (20.4) |  |
| Non-Hispanic/Latino | 1622 (59.3) | 979 (55.9) | 294 (64.6) | 349 (65.8) |  |
| Unknown Ethnicity | 352 (12.9) | 225 (12.8) | 54 (11.9) | 73 (13.8) |  |
| Age (%) |  |  |  |  | <0.001 |
| (18,30] | 49 ( 1.8) | 46 (2.6) | 2 ( 0.4) | 1 ( 0.2) |  |
| (30,40] | 161 ( 5.9) | 146 (8.3) | 3 ( 0.7) | 12 ( 2.3) |  |
| (40,50] | 248 ( 9.1) | 209 (11.9) | 21 ( 4.6) | 18 ( 3.4) |  |
| (50,60] | 470 (17.2) | 357 (20.4) | 62 (13.6) | 51 ( 9.6) |  |
| (60,70] | 694 (25.4) | 474 (27.1) | 114 (25.1) | 106 (20.0) |  |
| (70,80] | 596 (21.8) | 337 (19.2) | 117 (25.7) | 142 (26.8) |  |
| (80,90] | 400 (14.6) | 153 (8.7) | 104 (22.9) | 143 (27.0) |  |
| (90,100] | 117 ( 4.3) | 29 (1.7) | 32 ( 7.0) | 56 (10.6) |  |
| Comorbidities |  |  |  |  |  |
| Atrial fibrillation (N (%)) | 206 (7.5) | 91 ( 5.2) | 46 (10.1) | 69 (13.0) | <0.001 |
| Asthma (N (%)) | 229 (8.4) | 154 ( 8.8) | 36 ( 7.9) | 39 ( 7.4) | 0.537 |
| Coronary artery disease (N (%)) | 453 (16.6) | 171 ( 9.8) | 97 (21.3) | 185 (34.9) | <0.001 |
| History of cancer (N (%)) | 195 (7.1) | 123 ( 7.0) | 38 ( 8.4) | 34 ( 6.4) | 0.481 |
| Chronic kidney disease (N (%)) | 273 (10.0) | 90 ( 5.1) | 66 (14.5) | 117 (22.1) | <0.001 |
| COPD (N (%)) | 158 (5.8) | 70 ( 4.0) | 39 ( 8.6) | 49 ( 9.2) | <0.001 |
| Diabetes (N (%)) | 719 (26.3) | 378 (21.6) | 153 (33.6) | 188 (35.5) | <0.001 |
| Heart Failure (N (%)) | 276 (10.1) | 75 ( 4.3) | 67 (14.7) | 134 (25.3) | <0.001 |
| Hypertension (N (%)) | 1065 (38.9) | 595 (34.0) | 205 (45.1) | 265 (50.0) | <0.001 |
| **Laboratory Values** |  |  |  |  |  |
| Hemoglobin, G/dL (median [IQR]) | 12.70 [11.30, 13.90] | 12.90 [11.70, 14.00] | 12.60 [11.20, 13.90] | 11.90 [10.20, 13.40] | <0.001 |
| Lymphocyte (%) (median [IQR]) | 13.20 [8.12, 20.30] | 14.90 [9.10, 21.87] | 11.20 [7.10, 18.45] | 9.70 [6.20, 14.60] | <0.001 |
| D-dimer, ug/mL (median [IQR]) | 1.43 [0.79, 2.75] | 1.17 [0.71, 2.15] | 1.65 [1.05, 3.21] | 2.54 [1.51, 4.93] | <0.001 |
| D-dimer above 1 mcg/mL (N (%)) | 1453 (66.2) | 846 (58.5) | 282 (76.4) | 325 (85.8) | <0.001 |
| C-reactive protein, mg/L (median [IQR]) | 126.69 [63.71, 214.20] | 114.25 [56.61, 194.80] | 136.78 [72.30, 228.95] | 149.94 [95.09, 246.65] | <0.001 |
| Creatine kinase, U/L (median [IQR]) | 177.50 [83.25, 502.50] | 136.00 [72.50, 326.75] | 336.00 [120.00, 981.50] | 332.00 [155.00, 1015.00] | <0.001 |
| Lactate dehydrogenase, U/L (median [IQR]) | 441.00 [332.00, 592.00] | 425.00 [325.00, 551.00] | 456.00 [339.50, 616.75] | 520.00 [368.00, 753.50] | <0.001 |
| Ferritin, ng/mL (median [IQR]) | 780.50 [376.00, 1899.00] | 724.00 [350.75, 1629.75] | 828.00 [378.00, 1858.50] | 1093.00 [488.00, 2696.00] | <0.001 |
| Procalcitonin, ng/mL (median [IQR]) | 0.21 [0.09, 0.69] | 0.15 [0.07, 0.38] | 0.30 [0.12, 0.80] | 0.81 [0.28, 2.59] | <0.001 |
| Creatinine, mg/dL (median [IQR]) | 0.98 [0.75, 1.58] | 0.85 [0.70, 1.12] | 1.25 [0.90, 2.08] | 2.09 [1.24, 4.48] | <0.001 |
| Prothrombin time (PT), seconds (median [IQR]) | 14.30 [13.50, 15.60] | 14.00 [13.30, 14.90] | 14.40 [13.80, 16.48] | 15.20 [14.10, 16.80] | <0.001 |
| Partial thromboplastin time (aPTT), seconds (median [IQR]) | 32.90 [29.52, 37.90] | 32.30 [29.40, 36.80] | 33.00 [29.70, 38.98] | 34.40 [30.10, 40.70] | 0.002 |
| Albumin, G/dL (median [IQR]) | 3.00 [2.60, 3.30] | 3.00 [2.70, 3.30] | 2.90 [2.50, 3.20] | 2.90 [2.50, 3.20] | <0.001 |
| Bilirubin (Total), mg/dL (median [IQR]) | 0.60 [0.40, 0.80] | 0.60 [0.40, 0.80] | 0.60 [0.40, 0.80] | 0.60 [0.40, 0.90] | 0.048 |
| Sodium, mEq/L (median [IQR]) | 138.00 [135.00, 141.00] | 137.00 [135.00, 140.00] | 138.00 [135.00, 141.00] | 139.00 [136.00, 144.00] | <0.001 |
| Tachycardia (HR > 100 BPM) (N (%)) | 647 (23.6) | 393 (22.4) | 95 (20.9) | 159 (30.0) | <0.001 |
| Fever (>38.0°C) (N (%)) | 517 (18.9) | 372 (21.3) | 67 (14.8) | 78 (14.8) | <0.001 |
| Hypotension (SBP < 100 mmHg) (N (%)) | 228 (8.3) | 123 (7.0) | 40 ( 8.8) | 65 (12.3) | 0.001 |
| SBP above 160 mmHg (N (%)) | 227 ( 8.3) | 104 (5.9) | 53 (11.7) | 70 (13.2) | <0.001 |

Admission troponin-I concentrations are presented in **Figure 1**. Notably, 1751 (64%) patients had an initial troponin within the normal range. Few patients (86 patients, 3.1%) had an admission troponin elevation over 1 ng/mL within 24 hours of admission, while 173 (6.3%) had a troponin elevation over 1 ng/mL at any point during their hospital stay. Patient characteristics as well as admission vital signs and laboratory measurements, stratified by admission troponin-I, are also displayed in **Table 1**. Troponin elevations were categorized as mildly elevated and elevated as previously defined. Higher troponin concentrations were seen in patients who were over the age of 70.

**Figure 1.**
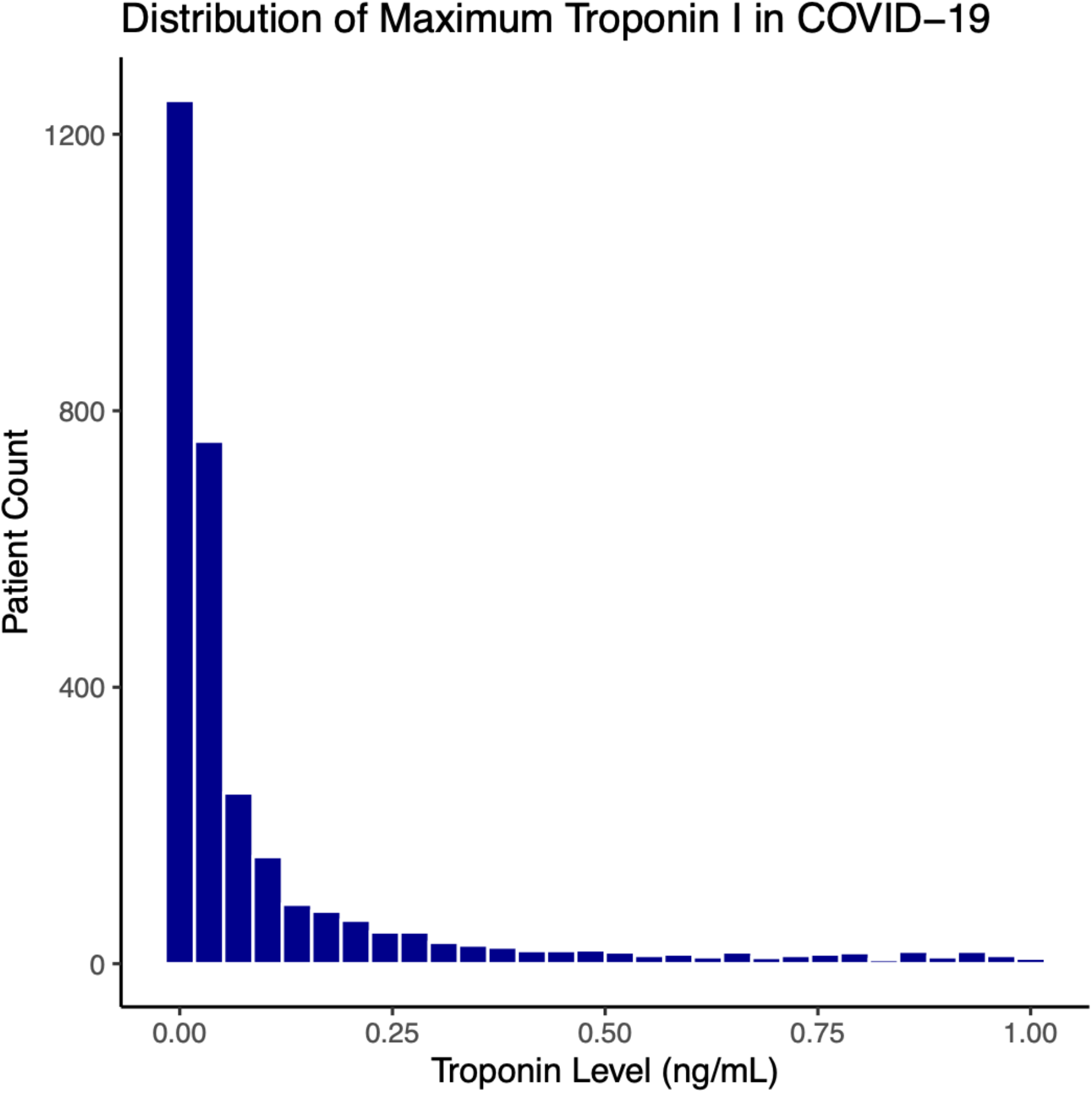
Distribution of maximum in-hospital troponin values for all patients with maximum troponin values below 1.0 ng/mL. Patients with troponin concentrations greater than 1.0 ng/mL are not shown.

The proportion of patients with CVD (defined here as CAD, AF, or HF) increased with higher troponin concentrations. Specifically, in those patients with significant myocardial injury (troponin I>0.09ng/mL), CVD including CAD, AF, and HF, was more prevalent (34.9%, 13.0%, and 25.3% respectively) compared to patients with mildly elevated troponins (21.3%, 10.1% and 14.7% respectively) and those with normal troponins (9.8%, 5.2%, and 4.3% respectively). **Figure 2** plots troponin measurements within 24 hours of admission among patients with CVD. Individuals with CVD generally presented with higher initial troponins than those without CVD. Similar trends for myocardial injury were seen in patients with history of HTN, DM, and CKD, but not in those with a history of asthma or cancer. Patient characteristics stratified by history of CVD, risk factors, and no history of either are shown in **Table 2**.

**Table 2.** Baseline characteristics of admitted patients stratified by history of cardiovascular disease, cardiovascular risk factors, or neither

| COVID-19 Patient Characteristics |  |  |  |  |  |
| --- | --- | --- | --- | --- | --- |
| Stratified by History of Cardiovascular Disease or Risk Factors, N = 2736 |  |  |  |  |  |
| Variable | All Patients | Cardiovascular Disease History |  |  | P Value |
|  |  | No Risk Factors | Risk Factors | Cardiovascular Disease |  |
| **Patient N (%)** | 2736 | 1374 (50.2) | 706 (25.8) | 656 (24.0) |  |
| **Sex (Female)** | 1106 (40.4) | 521 (37.9) | 309 (43.8) | 276 (42.1) | 0.022 |
| **Race (%)** |  |  |  |  | <0.001 |
| White | 634 (23.2) | 316 (23.0) | 130 (18.4) | 188 (28.7) |  |
| African American | 700 (25.6) | 323 (23.5) | 201 (28.5) | 176 (26.8) |  |
| Asian | 105 ( 3.8) | 48 ( 3.5) | 28 ( 4.0) | 29 ( 4.4) |  |
| Pacific Islander | 29 ( 1.1) | 8 ( 0.6) | 12 ( 1.7) | 9 ( 1.4) |  |
| Other | 1157 (42.3) | 602 (43.8) | 318 (45.0) | 237 (36.1) |  |
| Unknown Race | 111 ( 4.1) | 77 ( 5.6) | 17 ( 2.4) | 17 ( 2.6) |  |
| **Ethnicity (%)** |  |  |  |  | 0.001 |
| Hispanic/Latino | 762 (27.9) | 389 (28.3) | 214 (30.3) | 159 (24.2) |  |
| Non-Hispanic/Latino | 1622 (59.3) | 789 (57.4) | 401 (56.8) | 432 (65.9) |  |
| Unknown Ethnicity | 352 (12.9) | 196 (14.3) | 91 (12.9) | 65 ( 9.9) |  |
| **Laboratory Values** |  |  |  |  |  |
| Hemoglobin (g/dL) (median [IQR]) | 12.70 [11.30, 13.90] | 13.00 [11.80, 14.00] | 12.40 [11.00, 13.80] | 12.30 [10.47, 13.50] | <0.001 |
| Lymphocyte (%) (median [IQR]) | 13.20 [8.12, 20.30] | 13.10 [8.10, 20.10] | 13.90 [8.50, 21.20] | 12.90 [7.68, 19.72] | 0.133 |
| D-dimer ug/mL (median [IQR]) | 1.43 [0.79, 2.75] | 1.29 [0.73, 2.68] | 1.51 [0.85, 2.82] | 1.64 [0.91, 2.81] | <0.001 |
| D-dimer above 1 mcg/mL (N (%)) | 1453 (66.2) | 686 (61.9) | 410 (69.6) | 357 (71.8) | <0.001 |
| C-reactive protein, mg/L (median [IQR]) | 126.69 [63.71, 214.20] | 138.71 [72.36, 223.43] | 117.88 [61.33, 213.63] | 113.34 [53.74, 193.88] | <0.001 |
| Creatine kinase, U/L (median [IQR]) | 177.50 [83.25, 502.50] | 218.00 [93.00, 487.25] | 163.00 [70.75, 549.00] | 131.50 [79.00, 425.75] | 0.422 |
| Lactate dehydrogenase, U/L (median [IQR]) | 441.00 [332.00, 592.00] | 456.00 [351.25, 611.75] | 436.00 [329.00, 574.00] | 409.50 [305.00, 553.50] | <0.001 |
| Ferritin, ng/mL (median (IQR)) | 780.50 [376.00, 1899.00] | 814.00 [417.00, 1903.00] | 743.00 [356.00, 1850.00] | 765.00 [326.50, 1906.00] | 0.12 |
| Procalcitonin, ng/mL (median [IQR]) | 0.21 [0.09, 0.69] | 0.20 [0.09, 0.57] | 0.20 [0.09, 0.72] | 0.29 [0.10, 0.95] | <0.001 |
| Creatinine (median [IQR]) | 0.98 [0.75, 1.58] | 0.87 [0.70, 1.20] | 1.06 [0.78, 1.88] | 1.30 [0.90, 2.54] | <0.001 |
| Prothrombin time, seconds (PT) (median [IQR]) | 14.30 [13.50, 15.60] | 14.10 [13.50, 15.10] | 14.10 [13.30, 15.17] | 15.10 [13.90, 17.17] | <0.001 |
| Partial thromboplastin time, seconds (aPTT) (median [IQR]) | 32.90 [29.52, 37.90] | 32.30 [29.30, 36.20] | 32.00 [29.25, 36.55] | 35.70 [31.20, 42.27] | 0.002 |
| Albumin, g/dL (median [IQR]) | 3.00 [2.60, 3.30] | 3.00 [2.60, 3.30] | 3.00 [2.60, 3.30] | 3.00 [2.60, 3.30] | 0.703 |
| Bilirubin (Total), mg/dL (median [IQR]) | 0.60 [0.40, 0.80] | 0.60 [0.40, 0.80] | 0.50 [0.40, 0.70] | 0.60 [0.40, 0.90] | 0.002 |
| Sodium, mEq/L (median [IQR]) | 138.00 [135.00, 141.00] | 138.00 [135.00, 140.00] | 138.00 [135.00, 141.00] | 138.00 [135.00, 141.00] | 0.057 |
| Tachycardia (HR > 100 BPM) (N (%)) | 647 (23.6) | 357 (26.0) | 166 (23.5) | 124 (18.9) | 0.002 |
| Fever (>38.0°C) (N (%)) | 517 (18.9) | 283 (20.6) | 143 (20.3) | 91 (13.9) | 0.001 |
| Hypotension (SBP < 100 mmHg) (N (%)) | 228 (8.3) | 111 (8.1) | 58 (8.2) | 59 (9.0) | 0.762 |
| SBP above 160 mmHg (N (%)) | 227 ( 8.3) | 84 (6.1) | 68 (9.6) | 75 (11.5) | <0.001 |

**Figure 2.**
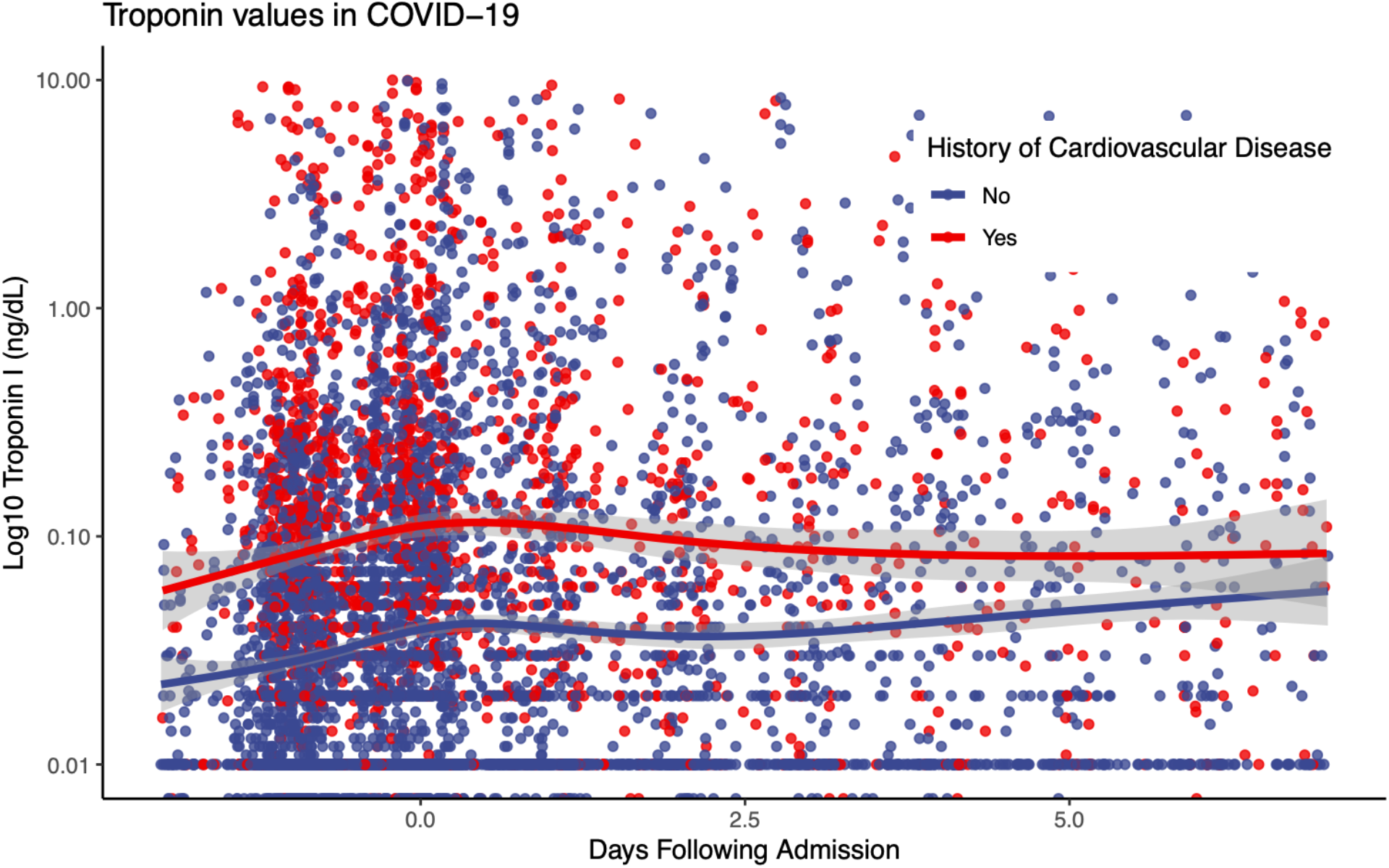
Plot of longitudinal troponin values over time, stratified by history of cardiovascular disease (CAD, HF, AFib) or no history of cardiovascular disease. Smoothing lines fit via LOESS regression with shaded areas indicating 95% confidence intervals.

Inflammatory markers were higher among patients with more substantial troponin elevations as well. In particular, the median D-dimer, C-reactive protein, lactate dehydrogenase, and procalcitonin were higher in patients with elevated initial troponins (2.54 ug/mL, 149.9 mg/L, 520.0 U/L, 0.81 ng/mL) than those with mildly elevated troponins (1.65 ug/mL, 136.78 mg/L, 456.0 U/L, 0.30 ng/mL) and those with normal troponins (1.17 ug/mL, 114.25 mg/L, 425.0 U/L, 0.15 ng/mL). Patients who had lower hemoglobin, hypo- or hypertension, or tachycardia generally presented with higher troponins than those who did not.

### Outcomes

Of 2736 COVID-19 patients included in our study, 506 (18.5%) died, 1132 (41.4%) were discharged, and 1098 (40.1%) remained hospitalized at the time of data freeze for this report. **Figure 3** presents cumulative incidence plots displaying probability for three possible outcomes (mortality, discharge from hospital, or continued hospitalization) over time. Milder forms of myocardial injury (e.g. troponin concentration 0.03-0.09 ng/mL) were associated with less frequent discharge and higher risk of death than troponin levels in the reference range after adjustment for clinically relevant covariates (adjusted HR: 1.77, 95% CI 1.39-2.26) (**Figure 4a)**. Troponin concentrations over 0.09 ng/dL were associated with more pronounced risk of death (adjusted HR 3.23, 95% CI 2.59-4.02) after adjustment. This risk was consistent across patients stratified by history of CVD, CVD risk factors such as DM or HTN only, and neither CVD nor risk factors. **(Figure 4b**). A sensitivity analysis using a competing-risks framework demonstrated similar adjusted hazards ratios for risk of death – HR 2.01 (95% CI: 1.59-2.54) for troponin concentrations >0.03-0.09 ng/mL, and HR 3.60 (95% CI: 2.88-4.52) for troponin concentrations >0.09 ng/mL.

**Figure 3.**
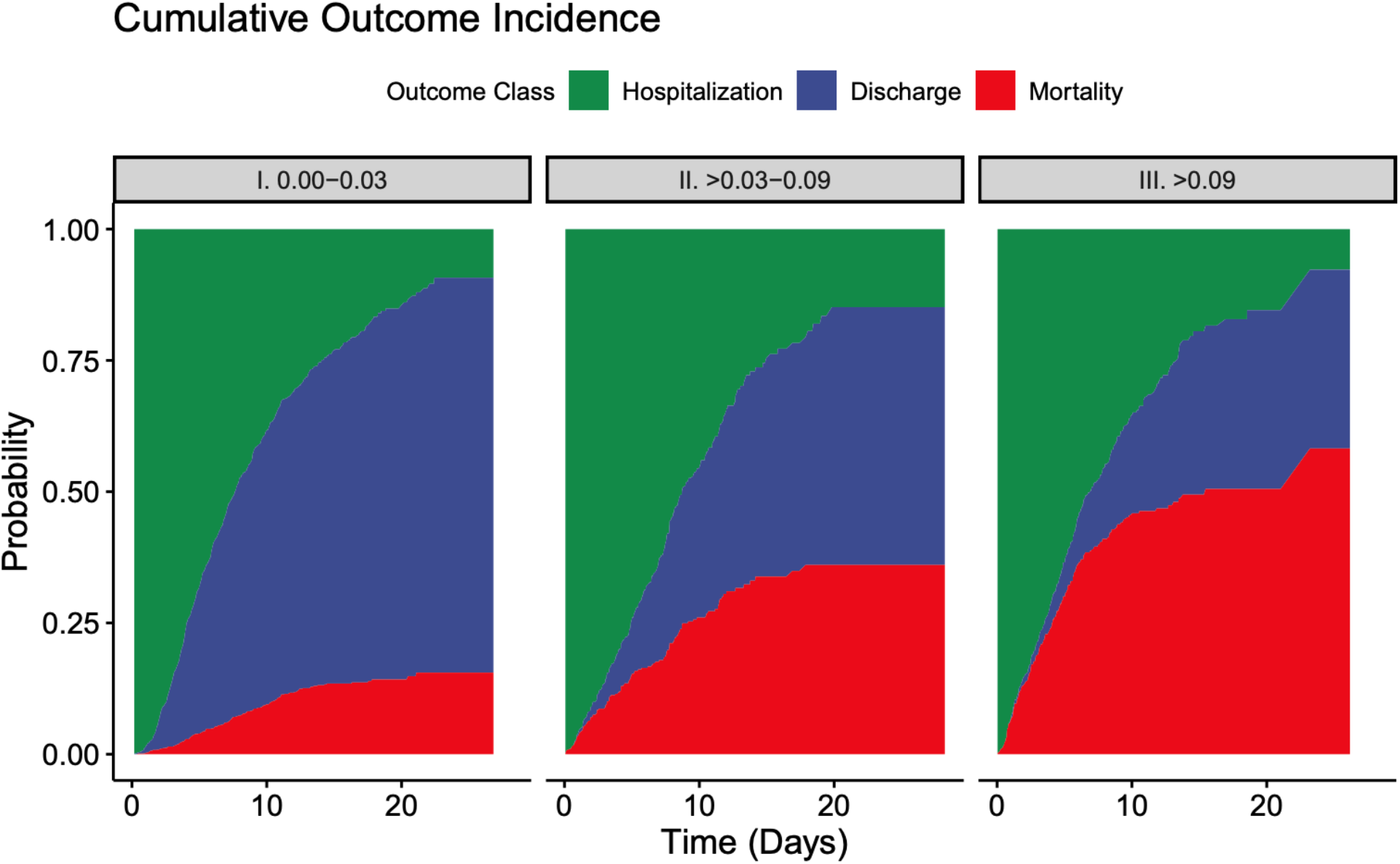
Cumulative incidence plots displaying probability for three possible outcomes (mortality, discharge from hospital, or continued hospitalization) over time.

**Figure 4a.**
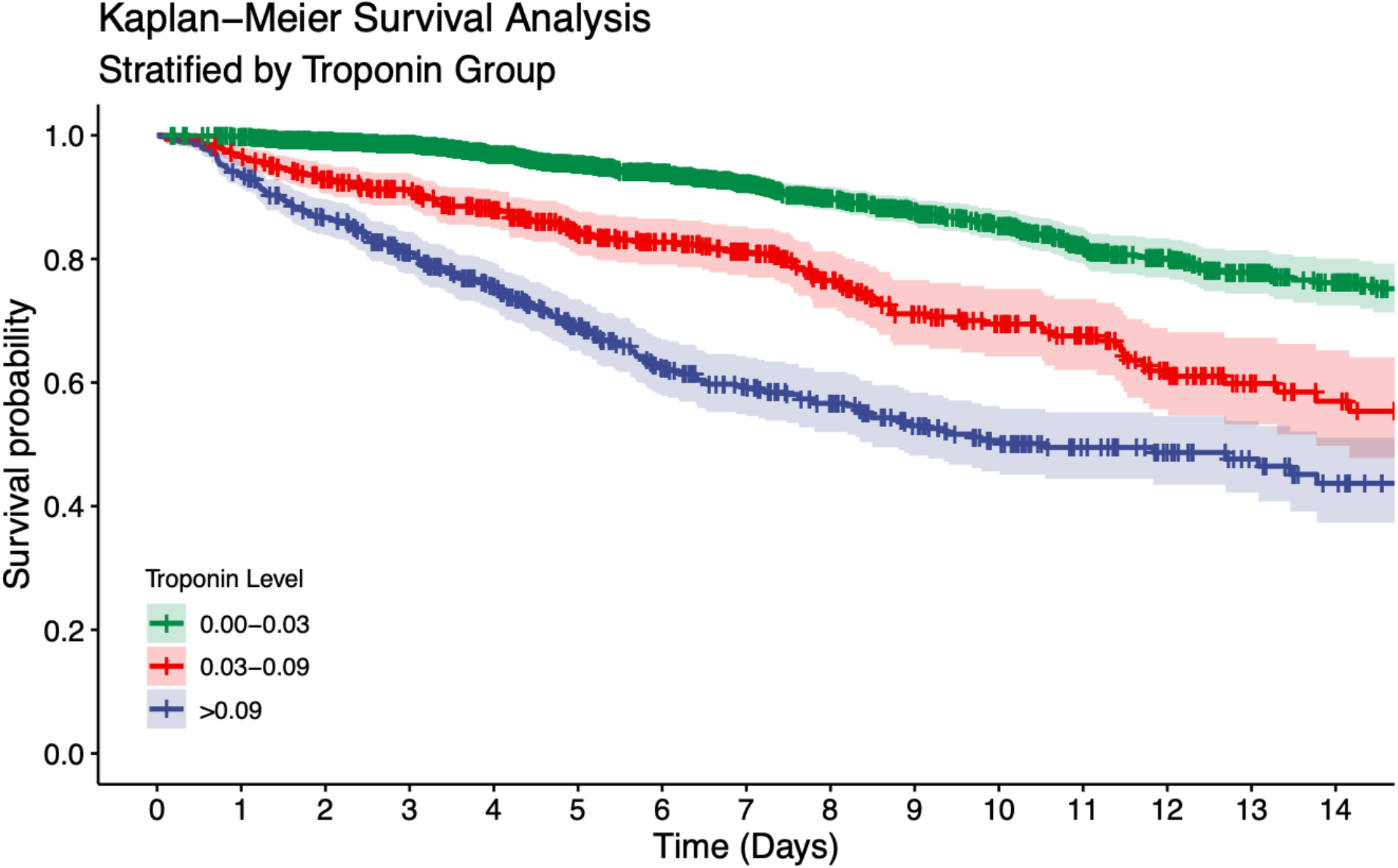
Kaplan-Meier plot for survival past hospital admission, stratified by troponin grouping. Patients were considered to be right-censored if they were discharged alive from the hospital or were still hospitalized at the time of data freeze (April 12, 2020). Survival times were significantly different between groups (p<0.001).

**Figure 4b.**
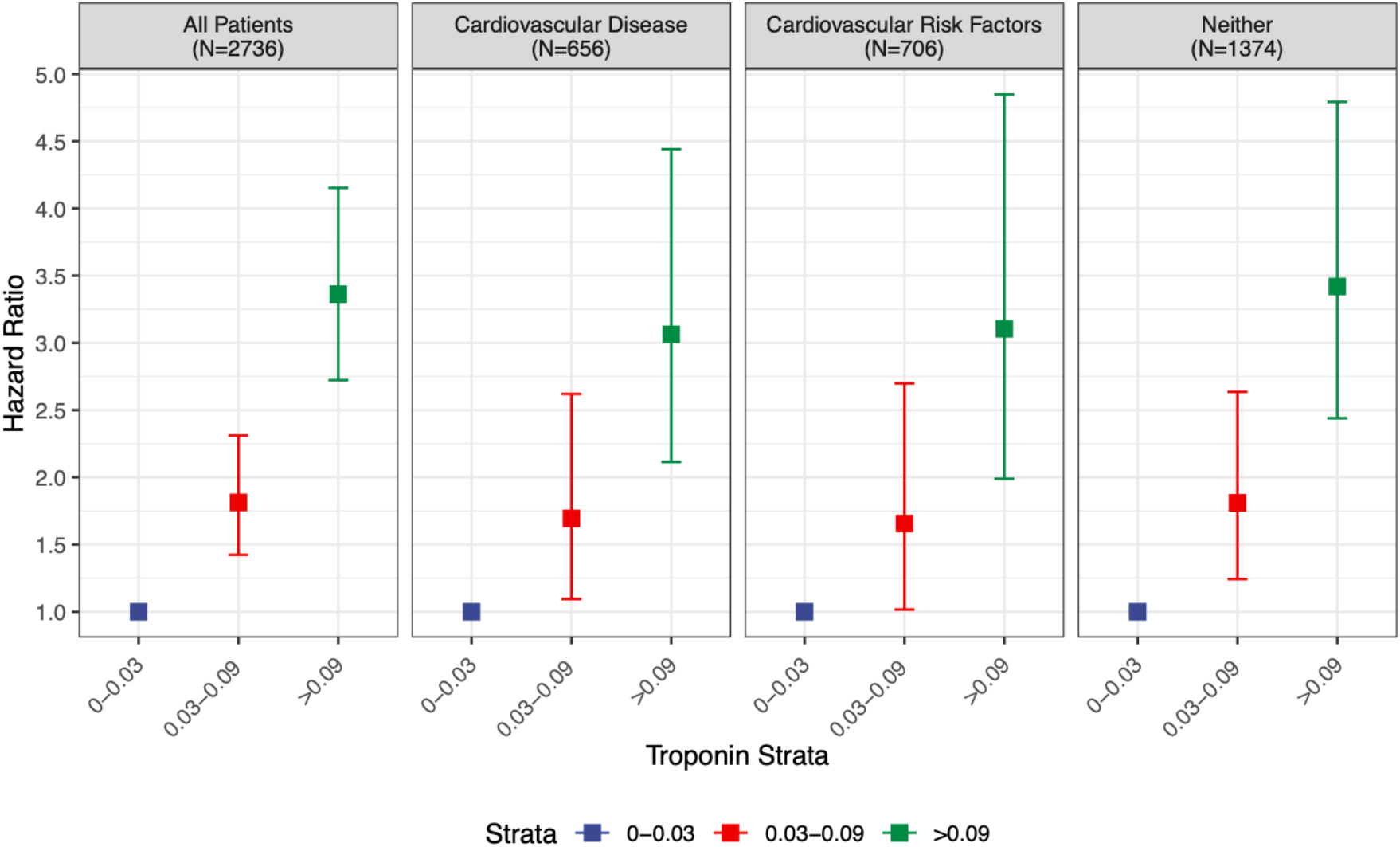
Hazard ratios and 95% confidence intervals calculated by Cox proportional hazards regression models for mortality stratified by comorbidities. Patients with cardiovascular disease had comorbidities of coronary artery disease, heart failure, or atrial fibrillation. Patients with cardiovascular risk factors had comorbidities of DM or HTN, but not cardiovascular disease

## DISCUSSION

Although pulmonary manifestations are the most common consequence, COVID-19 causes systemic inflammation with varying presentations of cardiac involvement as well (10). In this multihospital retrospective cohort study of nearly 3000 patients, we demonstrate the following observations: 1) Myocardial injury is common among patients hospitalized with COVID-19 but is more often mild, associated with low-level elevation in troponin concentration 2) More significant myocardial injury may be associated with more than a tripling in risk of mortality 3) COVID-19 patients with history of CVD are more likely to suffer myocardial injury than patients without CVD but without obvious corroborating evidence for primary acute myocardial infarction.

Though troponin elevation above the 99th percentile of the upper reference limit (URL) is considered the central marker of “myocardial injury” (11), underlying pathophysiologic mechanisms must be elucidated within the context of clinical circumstances. Myocardial injury is best recognized in the context of ischemia, however several non-ischemic mediated mechanisms, which include apoptosis, myocardial strain, myocyte necrosis, and increased cell membrane permeability mediated exocytotic release of troponin may contribute to such injury (12,13). Consequently, in assessing causal connections between COVID-19 and cardiac injury, it is critical to apply concepts described in the Fourth Universal Definition of Myocardial Infarction. In this regard, very few patients meet strict criteria for acute myocardial infarction; though some patients in this cohort certainly suffered ischemic myocardial damage from either Type 1 or 2 myocardial infarction, it is more likely the injury observed was mediated through a non-coronary mechanism. Challenges exist regarding understanding other potential mechanisms, however.

Despite several reports of COVID-19 associated myocarditis, to date, one case (14) demonstrated detection of SARS-CoV-2 viral particles in cardiac tissue but no case has demonstrated COVID-19 genome in cardiac tissue on biopsy or autopsy accompanied by troponin elevation consistent with criteria used to diagnose myocarditis (3,4,15-17). Other postulated mechanisms by which COVID-19 leads to cardiovascular morbidity include direct myocardial injury as a result of the inflammatory cascade or cytokine release, microvascular damage due to disseminated intravascular coagulation and thrombosis, direct entry of SARS-CoV-2 into myocardial cells by binding to ACE2 receptors, hypoxemia combined with increased metabolic demands of acute illness leading to myocardial injury akin to Type 2 Myocardial Infarction, and finally acute coronary syndrome from acute inflammation-triggered destabilization of atheromas (18-20). In a recent case series of 18 patients with COVID-19 infection and ST-segment elevation on electrocardiogram, 10 were deemed to have non-coronary myocardial injury by virtue of non-obstructive disease on coronary angiography and/or normal wall motion on echocardiography (21). Despite lower troponin concentrations in this group, 9 died as opposed to 4/8 in the ST-Elevation MI group, which may suggest higher mortality associated with non-ischemic mediated myocardial injury, however more data are needed.

In the present report, we demonstrate that myocardial injury was prevalent, occurring in 36% of hospitalized patients in the US. Evidence for myocardial injury was more frequent in our cohort compared to recent reports from China (2,22-25). These studies included between 41 and 416 patients and noted prevalence of myocardial injury ranging from 7-28%. Similar to these smaller reports, we also noted that patients with myocardial injury tended to be older and have a history of CVD. We also noted lower hemoglobin values, higher inflammatory markers, and more frequent rates of tachycardia or hypo/hypertension. The extent to which supply and demand imbalance is at play in the presence of underlying coronary disease is unknown, however overall these observations tend to support a non-ischemic mechanism of myocardial injury as opposed to dramatic primary acute coronary syndrome or myocarditis.

Despite being present as low-level concentrations, troponin elevation to greater than 3 times the URL was associated with a three-fold increased risk of mortality despite adjustment for clinically relevant factors. This finding is in keeping with a report from Wuhan, China of 416 patients by Shi et al, which demonstrated a hazard ratio of 3.41 [95% CI, 1.62-7.16] for death in patients with myocardial injury as compared to patients without. Guo and colleagues reported similar findings among 187 patients also in Wuhan but emphasized that although myocardial injury was more prevalent in patients with history of CVD, outcomes were more favorable in patients with CVD and no myocardial injury as compared to individuals with myocardial injury and no history of CVD. We similarly show that myocardial injury when present, regardless of history of CVD or risk factors was associated with worse outcomes.

### Limitations

There are some notable limitations of the present analysis. First, there are limitations inherent to the use of EHR for patient level data in such a large sample size not explicitly verified by manual chart review. Despite these limitations, the use of EHR enabled timely analysis and rapid dissemination of crucial information in a large patient cohort at the epicenter of the pandemic. Second, some patients included had not completed their hospital course at the time of data freeze. We accounted for this by conducting a secondary, complementary survival analysis where hospital discharge was treated as a competing risk as outlined in the Methods section. Results from our competing risks analysis were not meaningfully different from a standard survival analysis where discharged patients were simply considered to be right-censored. Lastly our outcomes analyses were focused upon troponin measurements made at hospital admission and less upon serial troponin measurements obtained over the course of each patient’s hospital stay, although we provide plots demonstrating trends in serial troponin measurements.

## CONCLUSION

Myocardial injury is prevalent among patients with acute COVID-19 and is associated with worse outcomes. Those with prior CVD are more likely to suffer myocardial injury related to COVID-19 infection. Troponin elevation among hospitalized patients likely reflects non-ischemic or secondary myocardial injury. These results suggest abnormal troponin concentrations on admission may be helpful with regard to triage decision-making. However, whether treatment strategies based on troponin concentrations would be expected to improve outcomes remains a testable hypothesis.

## PERSPECTIVES

### Clinical Competency in Medical Knowledge 1

Myocardial injury reflected by troponin elevation is common among patients hospitalized with COVID-19, most often present at low levels, and is associated with higher risk of mortality.

### Clinical Competency in Medical Knowledge 2

Patients with a history of cardiovascular disease are more likely to have myocardial injury than individuals with risk factors only and those without cardiovascular disease.

### Translational Outlook

Elevated troponin concentrations among patients hospitalized with COVID-19 likely represent non-ischemic or secondary myocardial injury.

## Data Availability

The Mount Sinai Institutional Review Board (IRB) approved this research under a regulatory protocol allowing for analysis of patient-level COVID-19 data.

## Abbreviations

AF: atrial fibrillation
CAD: coronary artery disease
CKD: chronic kidney disease
COPD: chronic obstructive pulmonary disease
COVID-19: coronavirus disease 2019
CVD: cardiovascular disease
DM: diabetes mellitus
HF: heart failure
HTN: hypertension
MSHS: Mount Sinai Health System
SARS-CoV-2: severe acute respiratory syndrome coronavirus-2

## Figure Legends

**Central Figure.**
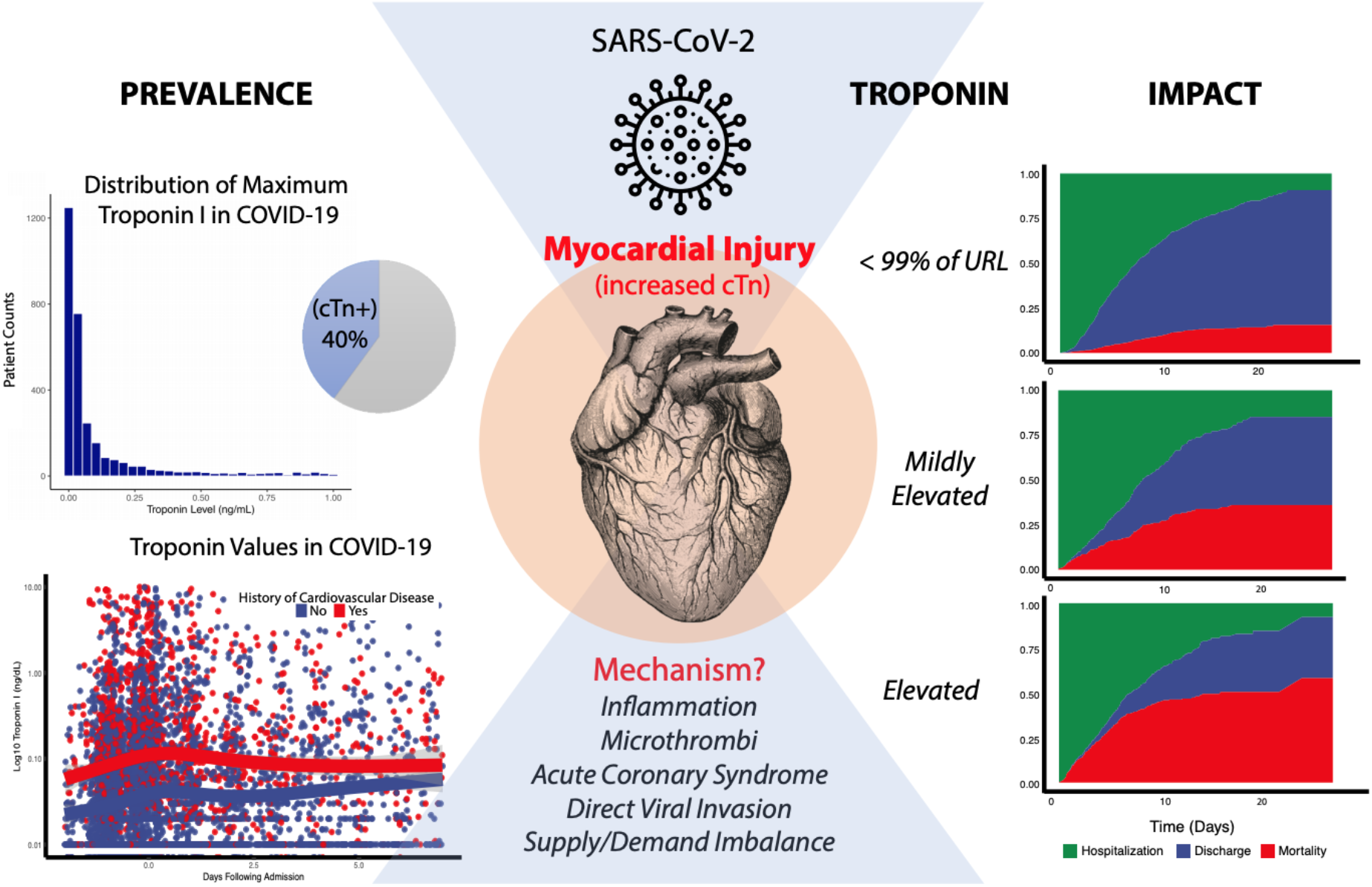
Myocardial injury reflected by troponin concentrations above the upper reference limit (URL) of 0.03ng/mL was present in 36 percent of patients hospitalized with COVID-19. Troponin levels among patients hospitalized with COVID-19 were generally under 1.0 ng/mL. Patients with a history of cardiovascular disease (in red, bottom left panel) including coronary artery disease, atrial fibrillation, and heart failure had higher troponin concentrations than patients without cardiovascular disease. Even small amounts of myocardial injury (e.g. troponin I 0.03-0.09ng/mL, n=455, 16.6%) were associated with death (adjusted HR: 1.77, 95% CI 1.39-2.26; P<0.001) while greater amounts (e.g. troponin I>0.09 ng/dL, n=530, 19.4%) were associated with more pronounced risk (adjusted HR 3.23, 95% CI 2.59-4.02). Troponin elevation in the setting of acute COVID-19 likely reflects non-ischemic or secondary myocardial injury.

## Acknowledgements

We would like to acknowledge Sayan Manna for his work in preparation of this manuscript.

